# Anxiety and Depression in Health Workers and General Population During COVID-19 Epidemic in IRAN: A Web-Based Cross-Sectional Study

**DOI:** 10.1101/2020.05.05.20089292

**Authors:** Leila Hassannia, Fatemeh Taghizadeh, Mahmood Moosazadeh, Mehran Zarghami, Hassan Taghizadeh, Azadeh Fathi Dooki, Mohammad Fathi, Reza Alizadeh Navaei, Akbar Hedayatizadeh-Omran

## Abstract

**Background:** The COVID-19 outbreak has exerted a great deal of psychological pressure on Iranian health workers and the general population. In the present study, the prevalence of anxiety and depression symptoms along with the related variables in this epidemic were investigated.

**Method:** An online cross-sectional study was conducted for the general public and healthcare workers in IRAN using a questionnaire comprised of demographic questions and Hospital Anxiety and Depression Scale. Chi square test was used to compare categorical variables, and univariate and multivariate logistic regression models were conducted.

**Results:** Of the 2045 participants,1136 (65.6%) were considered to have moderate and severe anxiety symptoms, and 865(42.3%) had moderate and severe depression symptoms. Based on the logistic regression models, the prevalence of anxiety was higher in the females than in the males (*OR*=1.4, 95% *CI:* 1.123-1.643, P=.002); the prevalence of anxiety was significantly higher in those aged 30-39 years than in other age groups (OR=1.6, 95% CI: 1.123-2.320, P=0.001); furthermore, the prevalence of anxiety and depression was significantly higher in doctors and nurses compared with other occupations (OR=1.9, 95% CI: 1.367-2.491, P< 0.001) and(OR=1.5, 95% CI: 1.154-2.021, P=0.003). In addition, the prevalence of anxiety symptoms in the likely-infected COVID-19 group was higher than in the noninfected COVID-19 group (OR=1.35, 95% CI: 1.093-1.654, P=0.005).

**Conclusions:** Regarding the high prevalence of anxiety and depression symptoms, especially among health care workers, appropriate psychological/psychiatric intervention necessitates.

## Introduction

COVID-19 is an acute respiratory illness with unknown causes that began in December 2019 in Wuhan, Hubei Province, China(1). In addition to causing physical injuries, this disease has seriously impacted the human mental health (2). On January 20, China confirmed the transfer of COVID-19 from human to human (3). Since then, people have shown behaviors associated with anxiety. Those in quarantined areas may experience boredom, anger, and loneliness. The symptoms of a viral infection such as cough and fever may cause cognitive distress and anxiety (4, 5). Several mental disorders such as depression, panic attack, delirium, suicidal thoughts, anxiety, and psychotic symptoms were reported during the early stages of SARS disease (4). During the COVID-19 epidemic and SARS disease in China, major psychological health issues were observed in young people, those spending a great amount of time on the disease, and healthcare workers(6). Due to the increase in the virus-related mortalities, both the medical staff and the general public have experienced psychological problems such as anxiety and depression (7). On average, the medical personnel work for 16 to 22 hours per day, putting their mental health at risk (8). High-risk positions with constant contact with the infected, high-risk infected environments with inadequate protection from contamination, lack of contact with family members, frustration, prejudice, loneliness, and fatigue further cause mental health issues such as stress, anxiety, depressive symptoms, insomnia, denial, anger, and fear (7). Mental health problems not only affect the medical staff’s attention, understanding, and decision-making ability, but may also prevent them from combatting the virus (7). Many of the medical and nursing staff working in Wuhan had mental health disorders (9). Medical staff not only tolerate the excess workload, but also run a high risk of infection. Furthermore, prolonged stress increases susceptibility to depression or other mental illnesses, leading to the increased risk of infection (10). Therefore, it is necessary to pay attention to protective factors and the process of successful adaptation to adverse conditions (11). The paucity of research in Iran, which is currently affected by this virus, has led researchers to investigate the effects of the pandemic on depression and anxiety in the general public and healthcare workers. Finally, it is important to establish early purposive mental health interventions throughout Iran.

## Settings and participants

The target population was the general Iranian population and health workers. Inclusion criteria were Iranian nationality, 15 years of age or more, willingness to participate, and ability to understand Persian language. Exclusion criteria were less than 15 years of age, major psychological disorders, and unwillingness to be studied. The project was conducted during quarantine from April 6 to April 15, 2020.

## Informed consent

Informed consent was obtained electronically prior to data collection. To ensure the quality of the survey, a response range was considered for items such as age which was restricted to 15-80 years, and the respondents were encouraged to carefully read the questionnaire instructions. Ultimately, a total of 2045 participants who completed the questionnaires were included in the data analysis.

## Investigation approach

The study was conducted during quarantine, including inter-provincial travel ban, prohibition of nonessential business activities, and closure of schools and universities.

Electronic “Porseline” Questionnaire was used as the survey tool and a professional online survey platform which can be used for the questionnaire survey, evaluation, and other purposes. Participants were recruited via advertisements on social media such as WhatsApp, Telegram, LinkedIn, and Instagram.

## Investigation tools

The present study made use of online demographic questionnaires and Hospital Anxiety and Depression Scale (HADS) for the general public and healthcare workers(8).

### 1- Demographic Information

a self-designed questionnaire was devised, comprising items related to the province of residence, age, gender, job, level of education, marital status, and an additional question about the infected individuals.

### 2- Hospital Anxiety and Depression Scale (HADS)

This questionnaire was developed by The questionnaire consists of two subscales, one related to depression and the other associated with anxiety (12, 13), and it includes a total of 14 questions. This scale is used to classify participants into three categories: without anxiety and/or depression (scores of 0 to 7), possible anxiety and/or depression (scores ranging from 8 to 10), and anxiety and/or depression (scores varying from 11 to 21) (14). This questionnaire was used on a general population of 6659 individuals aged 65-80 in Sweden; the internal consistency was 0.92 for the anxiety subscale and 0.88 regarding the depression subscale; moreover, the Cronbach’s alpha was 0.87 on the anxiety subscale and 0.81 on the depression scale(15). In Iran, the scale had an acceptable validity and reliability (Cronbach’s alpha of 0.78 and 0.86 on anxiety (HADS-S) and depression (HADS-D), respectively) (16, 17).

## Statistical Analysis

Descriptive analyses were performed to describe the demographic characteristics of the Iranian population. The prevalence of anxiety and depression symptoms, stratified by gender, age, education, occupation, and infection with COVID-19, was determined. Chi square test (*χ*^2^) was used to compare the categorical variables while independent sample t-test and ANOVA test compared the continuous variables with normal distribution. Univariate and multivariate logistic regression models were conducted to explore the potential contributing factors for anxiety and depression symptoms during COVID-19 outbreak. Odds ratio (*OR*) and 95% confidence interval were analyzed from logistic regression models. All data were analyzed using Statistical Package for Social Sciences (SPSS) version 24.0. *P*-values of less than 0.05 were considered as statistically significant (two-sided tests).

## Results

93% of the respondents completed the questionnaire on WhatsApp, 6% on Telegram, and 1% on other applications. The response rate was 59%, and the average time of response to questionnaire was 08.17 minutes. The results of this study are as follows:

### 1 General demographic data

Based on the online survey, qualified questionnaires were finally obtained The characteristics of the participants are shown in **Table 1**. Following the analysis of 2045 questionnaires, the average age of the respondents was 44.07 ± 11.638 (years), ranging from 20 to 60 years old, of whom 181 (8.9%) were under 30, 536 (26.2%) were 30-39 years old, 647 (31.6%) were 40-49 years old, 505 (24.7%) were 50-59 years old, and 176 (8.6%) were equal or above 60 years old. Of the 2045 participants, 127 (6.2%) were doctors, 105 (5.1%) were nurses, 229 (11.2%) were health staff, 26 (1.3%) were lab staff, and 580 (28.4%) were other employee staff. Moreover, 671 (32.8%) of the participants were males, 1374 (67.2%) were females, 1584 (77.5%) were married, 187 (11.8%) held a PhD, 494 (24.2%) had a MS, 738 (36.1%) had a MA, 110 (5.4%) had associate degrees, 330 (16.1%) had a high school diploma, and 132 (6.5%) had lower education levels.

**Table 1:**
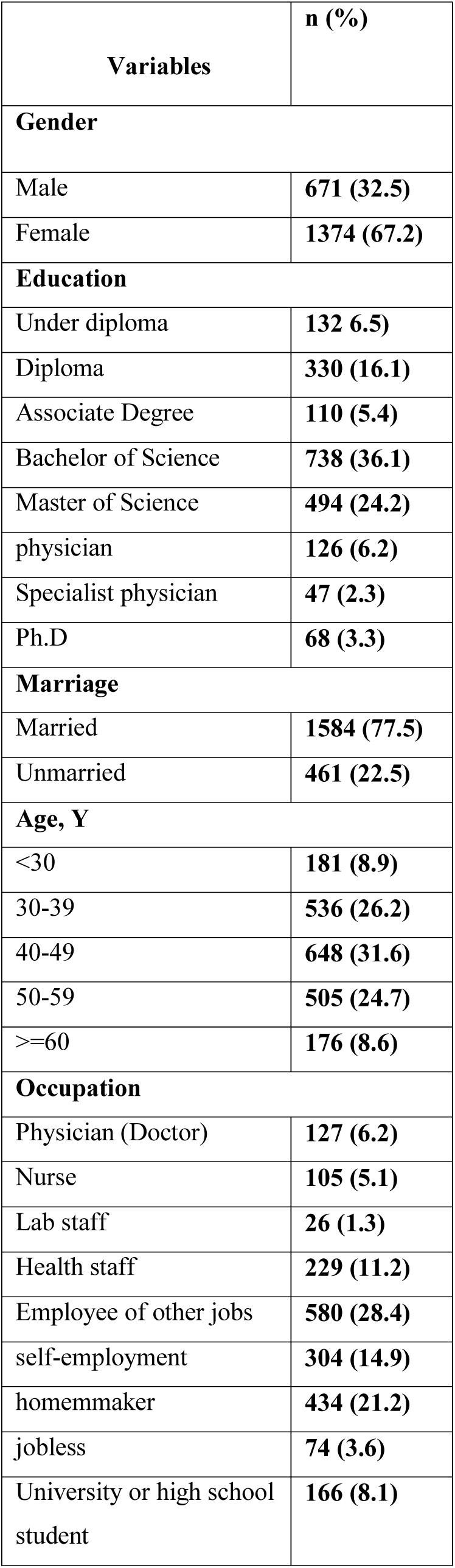
Demographic characteristics of study sample (N=2045)

### 2- Prevalence of anxiety and depression symptoms

**Tables 2 and 3** show the prevalence of anxiety and depression symptoms stratified by gender, education, marriage, age, and occupation. Univariate and multivariate logistic regression models are presented in **Tables 5 and 6**. Of the 2045 participants, 1136 (65.6%) had moderate and severe anxiety symptoms, and 865 (42.3%) had moderate and severe depression symptoms. According to **Tables 5 and 6** and the logistic regression models, the prevalence of anxiety in females was significantly higher than in males (*OR*=1.4, 95% *CI:* 1.123-1.643, P=.002). However, there were no significant differences between males and females concerning the prevalence of depression (*OR*=1.1, 95% *CI:* .912-1.336, P=.312). The prevalence of anxiety and depression in the age group 30-39 was significantly higher than in other age groups (OR=1.6, 95% CI: 1.123-2.320, P=0.001), (OR=1.17, 95% CI: .815-1.679, P=.394). The prevalence of anxiety and depression was significantly higher in doctors and nurses compared with other occupations (OR=1.9, 95% CI: 1.367-2.491, P< 0.001), (OR=1.5, 95% CI: 1.154-2.021, P=0.003). Other findings showed that the prevalence of anxiety and depression was not related to the marital status (P=.982, .354). Furthermore, the prevalence of anxiety in specialist physicians was significantly higher than in general physicians, and the anxiety of both groups was significantly higher than other education levels (X2=30.346, P=0.007); however, the prevalence of depression was not significant in education levels (X2=22.800, P=0.064).

**Table 2:** Prevalence of anxiety symptoms in participants during COVID-19 outbreak in Iran population stratified by gender, Education, Marriage, Age and, Occupation (N=2045)

| **Variables n (%)** | **Non-anxiety** | **Moderate-anxiety** | **Anxiety** | **X 2** | **p** |
| --- | --- | --- | --- | --- | --- |
| **Gender** |  |  |  |  |  |
| Male | **335 (49.9)** | **179(26.7)** | **157(23.4)** | **16.864** | **0.001** |
| Female | **574 (41.8)** | **368 (26.8)** | **432 (31.4)** |  |  |
| **Education** |  |  |  |  |  |
| Under diploma | **75 (56.8)** | **28 (21.2)** | **29 (22.0)** | **30.346** | **0.007** |
| Diploma | **150 (45.5)** | **87 (26.4)** | **93 (28.2)** |  |  |
| Associate Degree | **44 (40.0)** | **28 (25.5)** | **38 (24.5)** |  |  |
| Bachelor | **304 (41.2)** | **206 (27.9)** | **228 (30.9)** |  |  |
| Master of Science | **236 (47.8)** | **124 (25.1)** | **134 (27.1)** |  |  |
| General physician | **46 (36.5)** | **38 (30.2)** | **42 (33.3)** |  |  |
| Specialist physician | **15 (31.9)** | **16 (34)** | **16 (34.0)** |  |  |
| Ph.D | **39 (57.4)** | **20 (26.4)** | **9 (13.2)** |  |  |
| **Marriage** |  |  |  |  |  |
| Married | **932 (58.8)** | **329 (20.8)** | **323 (20.4)** | **3.723** | **0.155** |
| Unmarried | **248 (53.8)** | **107 (23.2)** | **106 (23)** |  |  |
| **Age, Y** |  |  |  |  |  |
| <30 | **92 (50.8)** | **43 (23.8)** | **46 (25.4)** | **45.486** | **0.001** |
| 30-39 | **204 (38.1)** | **142 (26.5)** | **190 (35.4)** |  |  |
| 40-49 | **263 (40.6)** | **181 (28)** | **203 (31.4)** |  |  |
| 50-59 | **245 (48.5)** | **138 (27.3)** | **122 (24.2)** |  |  |
| >=60 | **105 (59.7)** | **43 (24.4)** | **28 (15.9)** |  |  |
| **Occupation** |  |  |  |  |  |
| Physician (Doctor) | **40 (31.5)** | **42 (33.1)** | **45 (35.4)** | **34.846** | **0.004** |
| Nurse | **33 (31.4)** | **29 (27.6)** | **43 (41)** |  |  |
| Lab staff | **11 (42.3)** | **7 (26.9)** | **8 (30.8)** |  |  |
| Health staff | **97 (42.4)** | **61 (26.6)** | **71 (31.0)** |  |  |
| Employee of other jobs | **290 (50.0)** | **143 (24.7)** | **147 (25.3)** |  |  |
| self-employment | **132 (43.10)** | **78 (25.7)** | **94 (30.9)** |  |  |
| housewife | **187 (43.1)** | **126 (29.0)** | **121 (27.9)** |  |  |
| jobless | **30 (40.5)** | **20 (27.0)** | **24 (32.4)** |  |  |
| University or high school student | **89 (53.6)** | **41(24.7)** | **36 (21.7)** |  |  |

**Table 3:** Prevalence of depression symptoms in participants during COVID-19 outbreak in Iran population stratified by gender, Education, Marriage, Age and, Occupation (N=2045)

| **Variables n(%)** | **Non-depression (n=62)** | **Moderate-depression (n=62)** | **depression (n=24)** | **X 2** | **p** |
| --- | --- | --- | --- | --- | --- |
| **Gender** |  |  |  |  |  |
| Male | **403 (60.1)** | **138 (20.6)** | **130 (19.4)** | **2.453** | **0.293** |
| Female | **777 (56.6)** | **298 (21.7)** | **299 (21.8)** |  |  |
| **Education** |  |  |  |  |  |
| Under diploma | **79 (59.8)** | **31 (23.5)** | **22 (16.7)** | **22.800** | **0.064** |
| Diploma | **192 (58.2)** | **66 (20.0)** | **72 (21.8)** |  |  |
| Associate Degree | **56 (50.9)** | **24 (21.8)** | **30 (27.3)** |  |  |
| Bachelor | **418 (56.6)** | **156 (21.1)** | **164 (22.2)** |  |  |
| Master of Science | **307 (62.1)** | **97 (19.6)** | **90 (18.2)** |  |  |
| General physician | **60 (47.6)** | **37 (29.4)** | **29 (23.0)** |  |  |
| Specialist physician | **22 (46.8)** | **11 (23.4)** | **14 (29.8)** |  |  |
| Ph.D | **46(67.6)** | **14 (20.6)** | **8 (11.8)** |  |  |
| **Marriage** |  |  |  |  |  |
| Married | **932 (58.8)** | **329 (20.8)** | **323 (20.4)** | **3.723** | **0.155** |
| Unmarried | **248 (53.8)** | **107 (23.2)** | **106 (23.0)** |  |  |
| **Age, Y** |  |  |  |  |  |
| <30 | **101 (55.8)** | **40 (22.1)** | **40 (22.1)** | **26.098** | **0.001** |
| 30-39 | **280 (52.2)** | **129 (24.1)** | **127 (23.7)** |  |  |
| 40-49 | **369 (57.0)** | **127 (19.6)** | **151 (23.3)** |  |  |
| 50-59 | **309 (61.2)** | **115 (22.8)** | **81 (16.0)** |  |  |
| >=60 | **121 (68.8)** | **25 (14.2)** | **30 (17.0)** |  |  |
| **Occupation** |  |  |  |  |  |
| Physician (Doctor) | **61 (48.0)** | **32 (25.2)** | **34 (26.8)** | **28.165** | **0.30** |
| Nurse | **51 (48.6)** | **27 (25.7)** | **27 (25.7)** |  |  |
| Lab staff | **15 (57.7)** | **6 (23.1)** | **5 (19.2)** |  |  |
| Health staff | **125 (54.6)** | **49 (21.4)** | **55 (24.0)** |  |  |
| Employee of other jobs | **367 (63.3)** | **110 (19.0)** | **103 (17.8)** |  |  |
| self-employment | **176 (57.9)** | **61 (20.1)** | **67 (22.0)** |  |  |

| **Virus infection status** | **I was infected with the virus and was treated (Confirmed infected)** | **I was infected with the virus and being under the treatment (Confirmed infected)** | **I was likely infected with the virus and recovered with the symptoms of a cold (likely infected)** | **I have never been infected with this virus (No infected)** | **X 2** | **P** |
| --- | --- | --- | --- | --- | --- | --- |
| **Variables**<br>**n (%)** |  |  |  |  |  |  |

### 3- Prevalence of anxiety and depression symptoms in the COVID-19 infected

The results of the analysis of the infected are shown in **Table 4, 5, and 6**. The prevalence of anxiety symptoms in likely-infected COVID-19 group was higher than in the noninfected COVID-19 group (OR=1.35, 95% CI: 1.093-1.654, P=0.005), and compared to the infected group was not statically significant (OR=1.13, 95% CI:.734-1.744, P=.577). Meanwhile, the prevalence of depression symptoms in the likely-infected COVID-19 group was not statistically significant compared with the noninfected (OR=1.2, 95% CI: .988-1.483, P=.066) and infected (OR=1.46, 95% CI: .956-2.239, P=.080) groups.

**Table 4:**
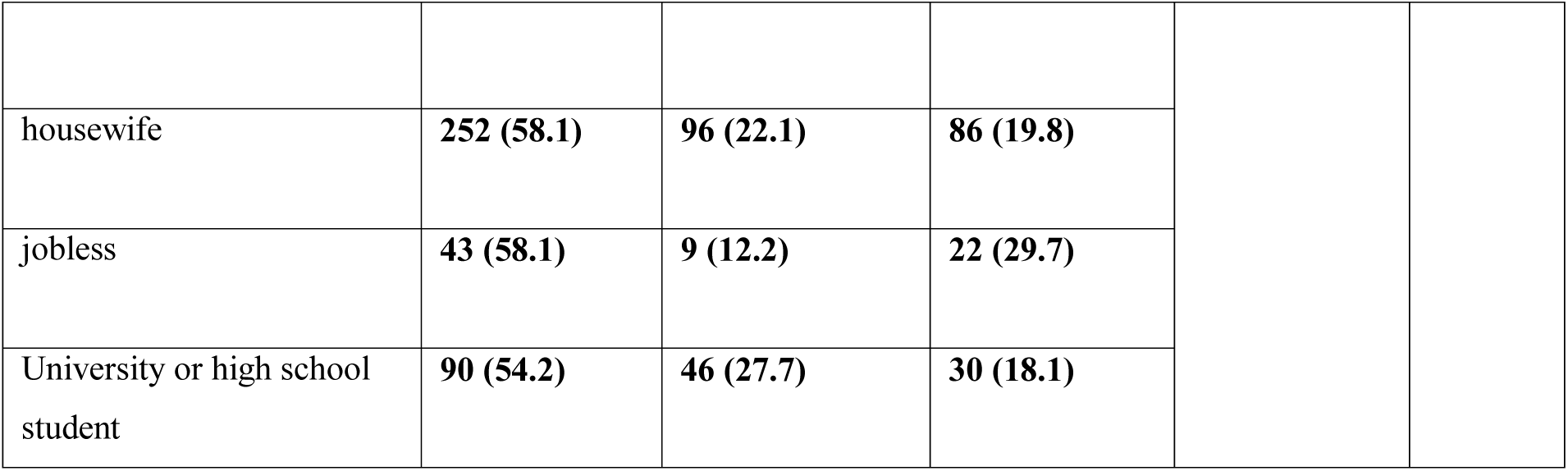
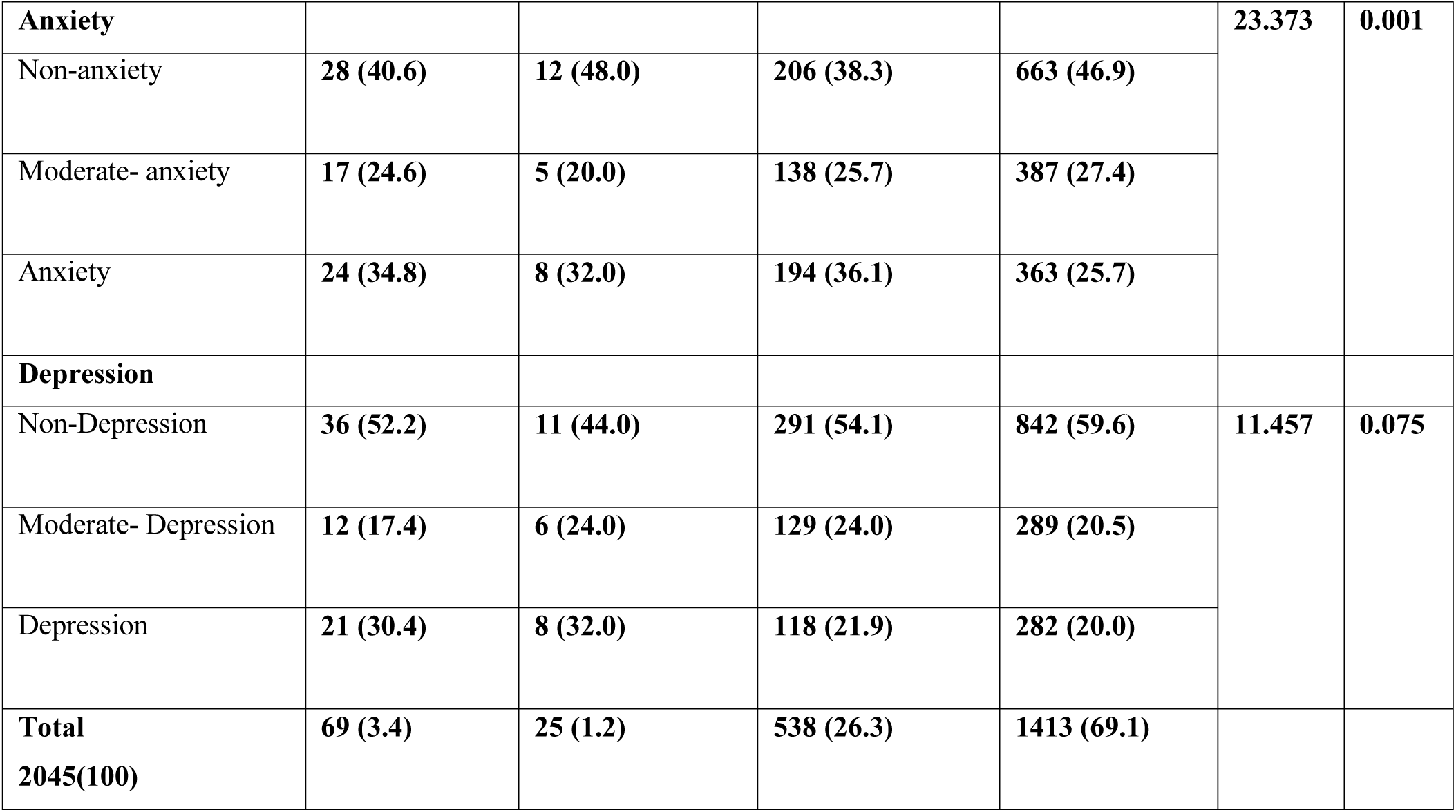
Prevalence of infected people and the relationship to anxiety and depression symptoms.

|  |  |  |  |
| --- | --- | --- | --- |
| housewife | **252 (58.1)** | **96 (22.1)** | **86 (19.8)** |
| jobless | **43 (58.1)** | **9 (12.2)** | **22 (29.7)** |
| University or high school student | **90 (54.2)** | **46 (27.7)** | **30 (18.1)** |

|  |  |  |  |  |  |  |
| --- | --- | --- | --- | --- | --- | --- |
| **Anxiety** |  |  |  |  | **23.373** | **0.001** |
| Non-anxiety | **28 (40.6)** | **12 (48.0)** | **206 (38.3)** | **663 (46.9)** |  |  |
| Moderate- anxiety | **17 (24.6)** | **5 (20.0)** | **138 (25.7)** | **387 (27.4)** |  |  |
| Anxiety | **24 (34.8)** | **8 (32.0)** | **194 (36.1)** | **363 (25.7)** |  |  |
| **Depression** |  |  |  |  |  |  |
| Non-Depression | **36 (52.2)** | **11 (44.0)** | **291 (54.1)** | **842 (59.6)** | **11.457** | **0.075** |
| Moderate- Depression | **12 (17.4)** | **6 (24.0)** | **129 (24.0)** | **289 (20.5)** |  |  |
| Depression | **21 (30.4)** | **8 (32.0)** | **118 (21.9)** | **282 (20.0)** |  |  |
| **Total**<br>**2045(100)** | **69 (3.4)** | **25 (1.2)** | **538 (26.3)** | **1413 (69.1)** |  |  |

**Table 5:** Results of univariate and multivariate logistic regression analyses of Anxiety symptoms in participants (N=2045)

| Variable |  | univariate |  |  | Multivariate |  |  |
| --- | --- | --- | --- | --- | --- | --- | --- |
|  |  | OR | CI 95% | P-value | OR | CI 95% | P-value |
| **Gender** | **Male** | **Ref.** | **Ref.** | **Ref.** | **Ref.** | **Ref.** | **Ref.** |
|  | **Female** | 1.4 | 1.154-1.673 | .001 | 1.4 | 1.123-1.643 | .002 |
| **Age group** | **<30** | **Ref.** | **Ref.** | **Ref.** | **Ref.** | **Ref.** | **Ref.** |
|  | **30-39** | 1.68 | 1.198-2.363 | .003 | 1.61 | 1.123-2.320 | .010 |
|  | **40-49** | 1.51 | 1.084-2.101 | .015 | 1.47 | 1.008-2.131 | .045 |
|  | **50-59** | 1.1 | .781-1.541 | .593 | 1.04 | .704-1.542 | .837 |
|  | **>=60** | .7 | .460-1.063 | .094 | .7 | .443-1.114 | .133 |
| **Marriage** | **Married** | **Ref.** | **Ref.** | **Ref.** | **Ref.** | **Ref.** | **Ref.** |
|  | **Non married** | 1.02 | .829-1.259 | .838 | .1 | .780-1.275 | .982 |
| **Jobs** | **Others** | **Ref.** | **Ref.** | **Ref.** | **Ref.** | **Ref.** | **Ref.** |
|  | **Doctors and nurses** | 1.86 | 1.392-2.496 | <0.001 | 1.85 | 1.367-2.491 | <0.001 |
| **Infected COVID-19** | **No infected** | **Ref.** | **Ref.** | **Ref.** | **Ref.** | **Ref.** | **Ref.** |
|  | **likely infected** | 1.43 | 1.163-1.745 | .001 | 1.35 | 1.093-1.654 | .005 |
|  | **Confirmed infected** | 1.19 | .783-1.820 | .412 | 1.13 | .734-1.744 | .577 |

**Table 6:** Results of univariate and multivariate logistic regression analyses of depression symptoms in participants (N=2045)

| **Variable** |  | **univariate** |  |  | **Multivariate** |  |  |
| --- | --- | --- | --- | --- | --- | --- | --- |
|  |  | **OR** | **CI 95%** | **P-value** | **OR** | **CI 95%** | **P-value** |
| **Gender** | **Male** | **Ref.** | **Ref.** | **Ref.** | **Ref.** | **Ref.** | **Ref.** |
|  | **Female** | 1.155 | .958-1.394 | .132 | 1.1 | .912-1.336 | .312 |
| **Age group** | **<30** | **Ref.** | **Ref.** | **Ref.** | **Ref.** | **Ref.** | **Ref.** |
|  | **30-39** | 1.154 | .823 -1.620 | .406 | 1.17 | .815-1.679 | .394 |
|  | **40-49** | .951 | .682-1.326 | .951 | .98 | .672-1.417 | .898 |
|  | **50-59** | .801 | .568-1.129 | .801 | .81 | .546-1.199 | .291 |
|  | **>=60** | .574 | .372-.885 | .574 | .61 | .378-.968 | .036 |
| **Married** | **Marriage** | **Ref.** | **Ref.** | **Ref.** | **Ref.** | **Ref.** | **Ref.** |
|  | **Non marriage** | 1.228 | .996-1.513 | .054 | 1.1 | .881-1.427 | .354 |
| **Jobs** | **Others** | **Ref.** | **Ref.** | **Ref.** | **Ref.** | **Ref.** | **Ref.** |
|  | **Doctors and nurses** | 1.536 | 1.168-2.020 | .002 | 1.53 | 1.154-2.021 | .003 |
| **Infected COVID-19** | **No infected** | **Ref.** | **Ref.** | **Ref.** | **Ref.** | **Ref.** | **Ref.** |
|  | **Likely infected** | 1.252 | 1.025-1.529 | .028 | 1.2 | .988-1.483 | .066 |
|  | **Confirmed infected** | 1.475 | .971-2.240 | .069 | 1.46 | .956-2.239 | .080 |

**Abbreviations:** *OR*, odds ratio; 95% *CI*, 95% confidence interval; COVID-19, 2019 Corona Virus Disease; Ref., Reference group

## Discussion

The aim of this study was to investigate the anxiety and depression symptoms in health workers and the general public during COVID-19 pandemic in Iran.

Based on the results of the current study, 65.6% of the participants had anxiety symptoms, and 42.3% had depression symptoms. Furthermore, women had more anxiety than men, and anxiety in those aged 30-39 was significantly higher than in other age groups. Anxiety and depression were significantly more prevalent in doctors and nurses compared with other occupations. Additionally, anxiety symptoms in the likely-infected COVID-19 group was higher than in the noninfected.

Previous studies reported emotional distress severity and symptoms of stress and anxiety in many people and health workers. The following are some of these studies:

### 1- Anxiety and depression symptoms in general population

Lee SM et al. made use of interviews and HADS and found that 11% of the participants were anxious, and 15.1% were depressed in the early stages of the outbreak (18). Cao et al. examined the psychological impacts of the COVID-19 pandemic on the students of a Chinese university; they reported that 0.9% of the students had severe anxiety, 21.3% had mild anxiety, and 2.7% had moderate anxiety(19). In the present study, 24.7% of the students had moderate anxiety, and 21.7% had depression symptoms. Guo et al.(20) reported a high prevalence of mental health problems in the general population during COVID-19. In a study on residual psychiatric disorders among the survivors of SARS, 25% of the patients were found to have PTSD symptoms while 15.6% had severe depression(5). Furthermore, Jeong H et al. found that 16.6% and 7.6% of the uninfected individuals in quarantine had feelings of anger and anxiety, respectively(21), whereas 53.1% of the uninfected participants had anxiety symptoms in the present study. Wang C et al. in a study on 1,210 respondents from 194 Chinese cities, 53.8% of the respondents rated the psychological impacts of the outbreak as moderate or severe. 16.5% reported moderate to severe depressive symptoms, and 28.8% reported moderate to severe anxiety symptoms. Females, students, and those with physical symptoms and poor health had significantly higher levels of anxiety and depression(22). This is in line with a systematic review where students, females, and individuals with the symptoms of COVID-19 had higher rates of depression and anxiety(23). 65.6% of the participants in our study had anxiety symptoms, and 42.3% had depression symptoms. The odds ratio of anxiety symptoms in females was 1.4 times more than that of male. Huang Y and Zhao N investigated 7,236 volunteers and reported that the overall prevalence of depressive symptoms in the general public was 20.1%. Moreover, young people had a significantly higher prevalence of depression symptoms in comparison with others (24). The multivariate logistic regression in our results revealed that the prevalence of depression symptoms in the age group >=60 years and the anxiety symptoms in those aged 30-39 years were higher than others.

### 2- Anxiety and depression symptoms in healthcare workers

The physicians showed higher scores of anxiety and depression compared with other medical staff (4). Mo Y et al. found that stress of 180 nurses taking care of COVID-19 patients was 39.91 ± 12.92 with a score of 39.91% (25). Guo et al. studied the psychological impact of COVID-19 pandemic on 11118 Chinese hospital staff. They observed that despite the insignificant psychological impact of COVID-19 on the medical staff in China, it was necessary to take measures to protect them from the adverse effects of COVID-19 (26). The results of SARS outbreak showed that 18%-57% of the treatment staff experienced emotional distress in the entire period of infection (27). Huang et al. studied the outbreak of generalized anxiety disorder and the depressive symptoms of COVID-19 on 7,236 Chinese people; their results revealed the significant impact of COVID-19 pandemic on the mental health of the participants(24).

Zhang et al. investigated the effects of psychiatric crisis intervention on the medical staff and patients’ families in COVID-19 hospitals. They reported that the intervention was conducive to reducing social anxiety during the pandemic(1). Previous studies reported that the mental health of medical workers was in a worse state compared with the general population (28–30), which is consistent with the present results.

Zhu J et al. found that the prevalence of anxiety and depression symptoms was 11.4% and 45.6% in physicians and 27.9% and 43.0% in nurses(31). In a study in Singapore and India, the depression symptoms was reported 10.6% and anxiety symptoms was %15.7 in medical workers(32).

In the current study, 51.4% and 52% of the nurses and doctors had depression symptoms while 68.6% and 68.5% had anxiety symptoms. The nurses and doctors in Iran have experienced higher symptoms of anxiety and depression than the chines(31) and other countries (32) during the outbreak. Perhaps, one of the reasons is the lack of sufficient personal protective equipment such as gloves and masks at the beginning of the outbreak due to the Iran economic sanctions.

Among the 994 medical and nursing staff in Wuhan, 36.9% had sub-threshold mental health disorders, 34.4% had mild disorders, 22.4% had moderate disorders, and 6.2% had severe disorders (9). Huang Y and Zhao N found that compared with other occupational groups, healthcare workers were likely to have a lower quality of sleep (24).

Zandifar and Badrfam (33) stated that the severity, uncertainty, and unpredictability of COVID-19, social quarantine, and misinformation contributed to anxiety. They further highlighted the need for both mental health services, particularly for vulnerable populations, and the establishment of social wealth to reduce the adverse psychological effects of the outbreak.

### 3- Anxiety and depression symptoms in infected COVID-19 people

Anxiety symptoms in infected COVID-19 likelihood was higher than the no infected COVID-19 group and, comparing to the confirmed infected group was not statically significant. While, depression symptoms in infected COVID-19 likelihood was not statically significant comparing to the no infected and infected groups. Nguyen HC et al. found that the people with Suspected COVID-19 symptoms had a higher depression likelihood than those without COVID-19(34).

## Conclusions and recommendations

The majority of the medical staff and general population in our study experienced anxiety and depression symptoms. To address the concerns about COVID-19 in Iran, a free 1480 hotline has been provided, mental health professionals are providing guidelines in hospitals and medical centers, and NGO groups consisting of psychologists are counselling for free. Healthcare providers can also offer suggestions for coping with anxiety and depression and link patients to social and mental health services. Since media reports can be emotionally distressing, COVID-related news should be checked and restricted.

Researchers should also assess the effects of COVID-19 on other vulnerable people, including children and adolescents, those residing in rural areas and lacking access to healthcare, and individuals belonging to lower socio-economic states. It is also necessary to develop mental health interventions which are time-limited, sensitive to culture, and can be taught to volunteers and healthcare workers.

## Limitations

In this cross-sectional study, we were not able to establish a causal link; furthermore, we used the self-rating scale to evaluate the depression and anxiety symptoms of the medical staff and the general public. Finally, the age group did not include those above 60 years of age possibly due to the lack of access to Internet smartphones, or applications.

## Data Availability

Data is available as request

## Compliance with Ethical Standards

### Ethical Consideration

All procedures were conducted according to the Declaration of Helsinki and was approved by the Ethics Committee of Mazandaran University of Medical Sciences. Electronic informed consent was obtained from all participants. They could leave without any explanation before the research All procedures conducted were approved by the Ethics Committee of Mazandaran University of Medical Sciences (IR. MAZUMS. REC.1399.7360).

### Author statement contributors

FT conceptualized and designed the study, review and revised the manuscript, and approved the final manuscript as submitted. LH, RA and AH designed the data collection instruments, coordinated and supervised data collection, carried out the initial analyses, and interpreted the data, drafted the initial manuscript, and approved the final manuscript as submitted. HT, MF, FF and MZ agree to be cooperation for all aspects of the study. All of the authors edited and approved the manuscript.

## Acknowledgments

The authors are grateful to Mazandaran University of Medical Sciences, all the respondents who participated in this study, and the research assistants and colleagues who kindly cooperated in the conduct of the study.

